# Antimicrobial Peptides and Systemic Inflammation: A Network Analysis

**DOI:** 10.64898/2025.12.26.25343039

**Authors:** Fabiano Pinheiro da Silva

## Abstract

Antimicrobial peptides (AMPs) are essential components of the innate immune system, exhibiting diverse mechanisms of action. This study investigates the roles of cathelicidin (LL-37), alpha-defensins, and the S100 proteins S100A8 and S100A9 in systemic inflammation associated with sepsis, severe COVID-19, and acute pancreatitis using whole-blood bulk RNA-sequencing data. Gene co-expression network analysis revealed that during septic shock and severe COVID-19, cathelicidin and alpha-defensins act synergistically in innate immune responses, while S100A8 and S100A9 function through distinct pathways related to mitochondrial metabolism and ubiquitin ligase binding. In contrast, the acute pancreatitis network displayed a different pattern, with *CAMP* co-expressed alongside *S100A8* and *S100A9*, whereas alpha-defensins were downregulated and associated with inhibited mucosal immune responses. These findings suggest that antimicrobial peptides contribute variably to systemic inflammation depending on the underlying insult, underscoring their complex, context-dependent roles in critical illness.

## 1. Introduction

Antimicrobial peptides (AMPs) are a large family of natural, synthetic, and predicted molecules, comprising more than 5,000 compounds with structural and functional diversity (https://aps.unmc.edu). They represent an evolutionarily conserved defense mechanism and are produced by bacteria, archaea, protists, fungi, plants, and animals.

AMPs are multifunctional molecules, possessing the ability to kill microbes directly as well as to regulate several arms of the immune response [1]. Their activity is complex, and many aspects remain unclear. Over the past decades, it has become evident that they not only play a role in immunity but also participate in several other cellular responses, such as wound healing, cell death, and even carcinogenesis.

In mammals, cathelicidins and defensins are the most extensively studied antimicrobial peptides. Cathelicidins have only one member in humans, named LL-37, while, based on disulfide topology, six alpha-defensins (human Neutrophil Peptides 1 to 4 and human Enteric Defensins 5 and 6) [2] and more than 10 beta-defensins have been described [3, 4], although many beta-defensins are detected mainly in the male genital tract.

S100 proteins constitute a family of 25 known small proteins, expressed only in vertebrates, that also play a prominent role in the regulation of inflammation and immunity. They are involved in several cellular processes, and some, such as S100A7, S100A12, and S100A15, are classified as antimicrobial peptides. The S100A8 and S100A9 proteins, however, are the most extensively investigated, demonstrating a crucial role in infection and immunity [5, 6]. S100A8 and S100A9 are secreted by neutrophils and monocytes into the circulation and tend to form a heterodimer known as calprotectin, its most stable form. Calprotectin binds calcium and other divalent metals, such as zinc, and exerts several intra- and extracellular biological functions.

Systemic inflammation is a crucial defense mechanism but can become excessive or prolonged in some diseases, leading to deleterious effects. Investigating the pathways that trigger and sustain systemic inflammation is essential to understanding the pathophysiology of critical illnesses. We believe antimicrobial peptides play an important role in this context. Here, using RNA-sequencing (RNA-Seq) data, we investigate the role of LL-37 and alpha-Defensins 1 to 4 in critically ill patients with sepsis, COVID-19, and acute pancreatitis. Due to their similarity with antimicrobial peptides, S100A8 and S100A9 were also evaluated. Since beta-defensins are poorly expressed in the blood, they were not investigated.

## 2. Patients and Methods

Whole-blood bulk RNA-Seq datasets were obtained from GEO, a public data repository of the United States National Institutes of Health (https://www.ncbi.nlm.nih.gov/geo). The septic shock samples were obtained from series GSE154918 [7]. Sixteen samples from septic shock patients and twelve samples from healthy volunteers were selected to achieve the same male-to-female ratio (50%) in both groups. The COVID-19 samples were obtained from series GSE179850 [8]. Thirty-one samples from severe COVID-19 patients and sixteen samples from healthy volunteers were selected to achieve the same male-to-female ratio (94%) in both groups. The acute pancreatitis samples were obtained from series GSE194331 [9]. Ten samples from severe acute pancreatitis patients and ten samples from healthy volunteers were selected to achieve the same male-to-female ratio (80%) in both groups.

Gene co-expression analyses were performed using BioNERO [10]. The modules detected were then analyzed via the EnrichR R interface (https://cran.r-project.org/web/packages/enrichR), using the Reactome and Gene Ontology (Biological Process and Molecular Function) databases, to investigate the molecular pathways associated with these genes. All analyses were performed using the R software for statistical computing (https://www.r-project.org).

## 3. Results

Enrichment analysis using tools like EnrichR plays a crucial role in interpreting large-scale omics data by identifying biological functions, pathways, and processes overrepresented within specific gene modules. In our study, we utilized Enrichr to analyze gene sets from each module, which allowed us to uncover significant biological themes that are likely relevant to the disease states examined.

Moreover, module-trait correlations offer a statistical framework to link gene expression modules with clinical phenotypes or disease states. By calculating the Pearson correlation coefficients between module eigengenes (summary expression profiles of modules) and disease traits, we identified which modules are significantly associated with the severity or presence of disease.

Together, enrichment analysis contextualizes the gene modules within known biological pathways and functions, enhancing our understanding of disease mechanisms. When combined with module-trait correlations, this approach helps prioritize specific gene networks for further investigation, potentially revealing novel therapeutic targets or biomarkers. This integrative strategy thus provides a comprehensive framework for translating complex omics data into meaningful biological insights.

The sepsis network inferred by BioNERO contains 5,000 nodes and 12,497,500 edges. *CAMP*, the LL-37 gene, and *DEFA1* to *DEFA4* (the alpha-Defensins 1 to 4 genes) were detected in module blue, while *S100A8* and *S100A9* were detected in module brown. Module blue contains 30 genes, and module brown contains 414.

The COVID-19 network contains 1,000 nodes and 499,500 edges. *CAMP* and *DEFA1* to *DEFA4* were detected in module greenyellow, while *S100A8* and *S100A9* were detected in module black. Module greenyellow contains 116 genes, and module black contains 638.

The acute pancreatitis network contains 800 nodes and 319,600 edges. *DEFA1* to *DEFA3* were detected in module blue. *DEFA4* was not detected. *CAMP*, *S100A8*, and *S100A9* were detected in module brown. Module blue contains 39 genes, and module brown contains 500.

**Table 1** lists the module in which each gene was detected in the sepsis, COVID-19, and acute pancreatitis networks and the strength of each gene. In weighted networks, the degree of a node, called strength, is measured as the sum of the weights of the edges containing the node.

**Table 1.**
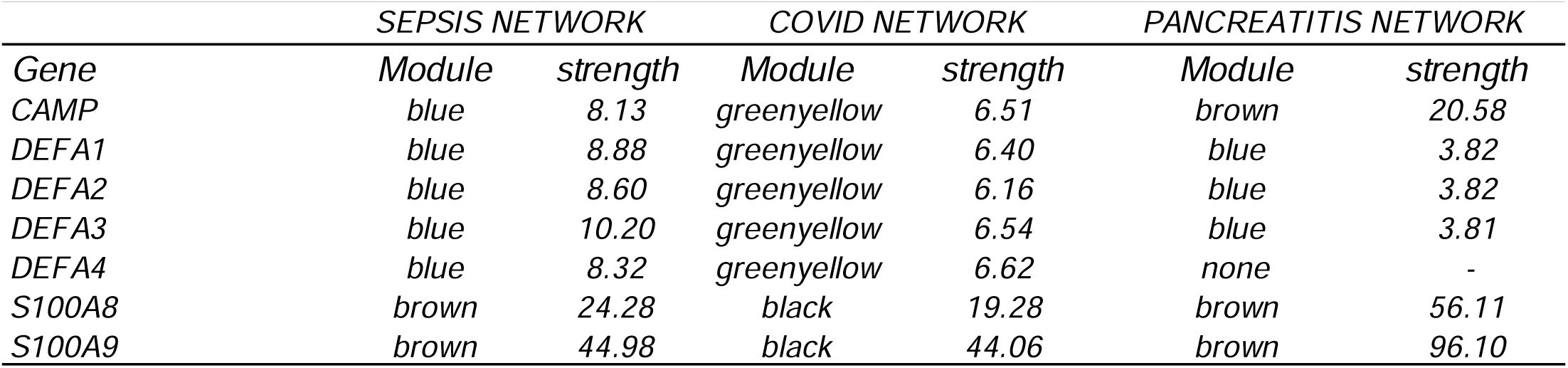
List of genes investigated, their respective module and strength in the septic shock, COVID-19 and acute pancreatitis networks.

### Septic patients gene co-expression analysis reveals that antimicrobial peptides and calprotectin act in different cell compartments

**Figure 1A** exhibits the module-trait correlations of the sepsis network. We can see that both module blue and brown are positively correlated with the sepsis state (Pearson coefficient equals 0.45 and 0.79, respectively). The expression profile for module blue is shown in **Figure 1B** and the one for module brown in **Figure 1C**.

**Figure 1.**
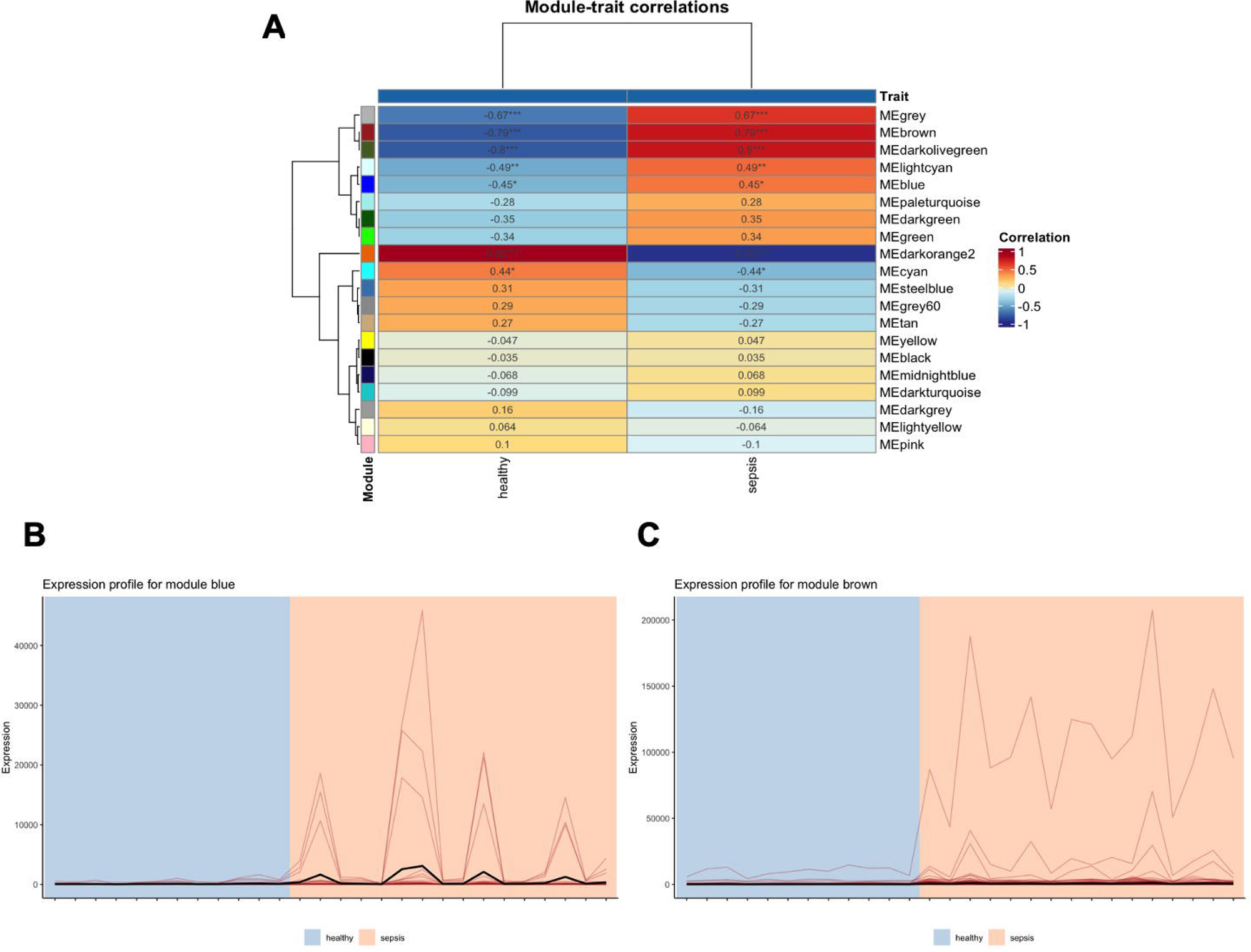
Module-trait relationship analysis of the sepsis network. (A) Correlation heatmap displaying the association between module eigengenes (MEs) and sepsis status; positive correlations are indicated by warm colors. Expression profiles of (B) module blue and (C) module brown are shown as line plots representing the gene expression patterns across samples. ME denotes module eigengenes summarizing the expression of genes within each module.

Regarding module blue, which contains *CAMP* and *DEFA1* to *DEFA4*, the Reactome database detected that these genes are associated with neutrophil degranulation, extracellular matrix organization and immunity (**Figure 2A**), while the Gene ontology database indicates participation in bacterial defense biological processes (**Figure 2B**) and molecular functions such as serine-type endopeptidase activity, calcium-dependent phospholipid binding, endopeptidase activity and protein kinase activity (**Figure 2C**).

**Figure 2.**
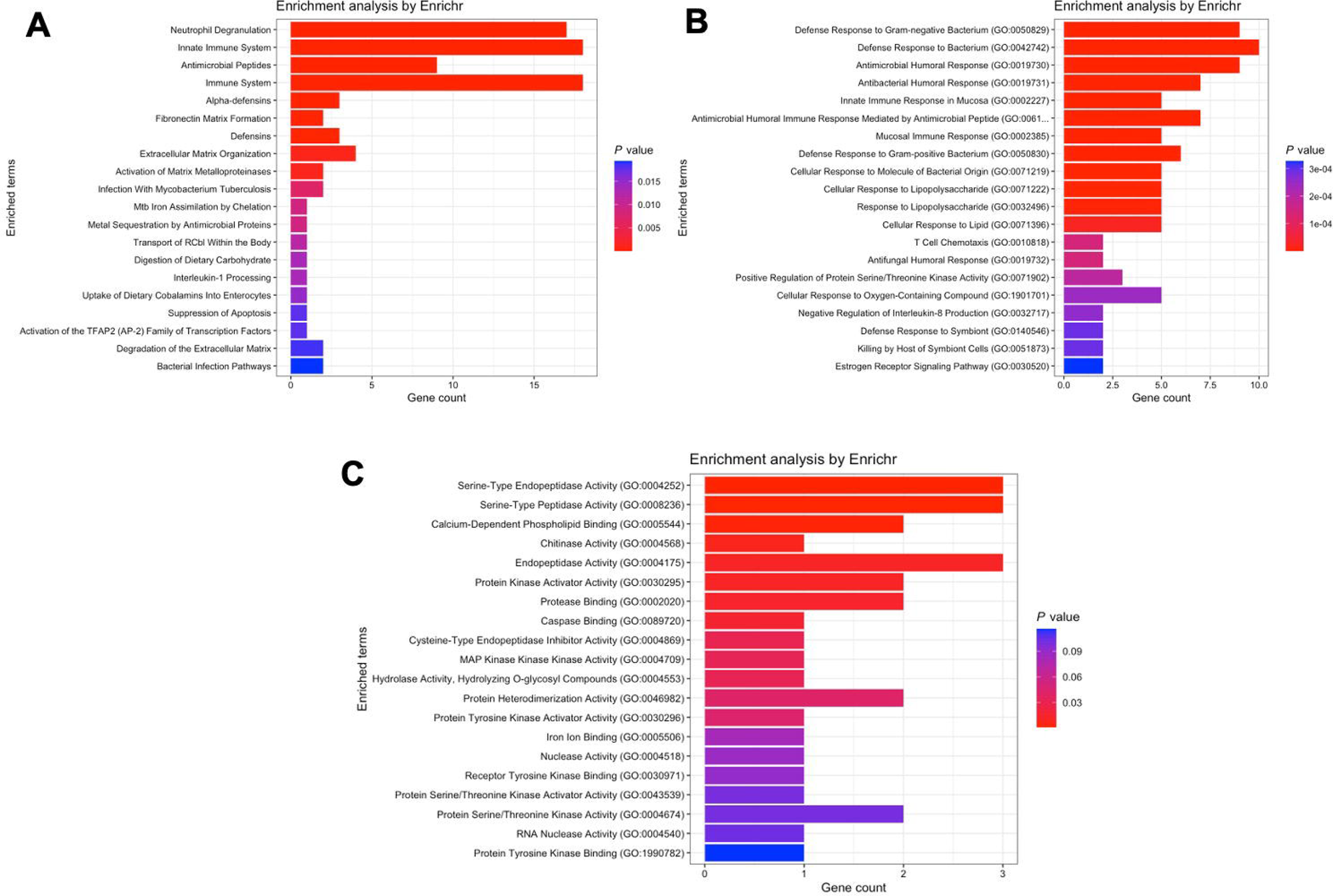
Pathway enrichment analysis for genes within module blue of the septic shock network. (A) Reactome pathway analysis highlighting biological pathways such as neutrophil degranulation, extracellular matrix organization, and immune responses. (B) Gene Ontology (GO) biological process terms associated with module blue, emphasizing bacterial defense mechanisms. (C) GO molecular functions enriched in this module, including serine-type endopeptidase activity, calcium-dependent phospholipid binding, endopeptidase activity, and protein kinase activity.

Analysis of module brown, which contains *S100A8* and *S100A9*, according to the Reactome database, suggests participation of these genes in immune responses, metabolism and respiratory electron transport (**Figure 3A**). The gene ontology databases, in the same direction, propose a role in biological processes such as aerobic respiration and neutrophil chemotaxis (**Figure 3B**) and in molecular functions such as ubiquitin ligase binding and metal ion binding (**Figure 3C**).

**Figure 3.**
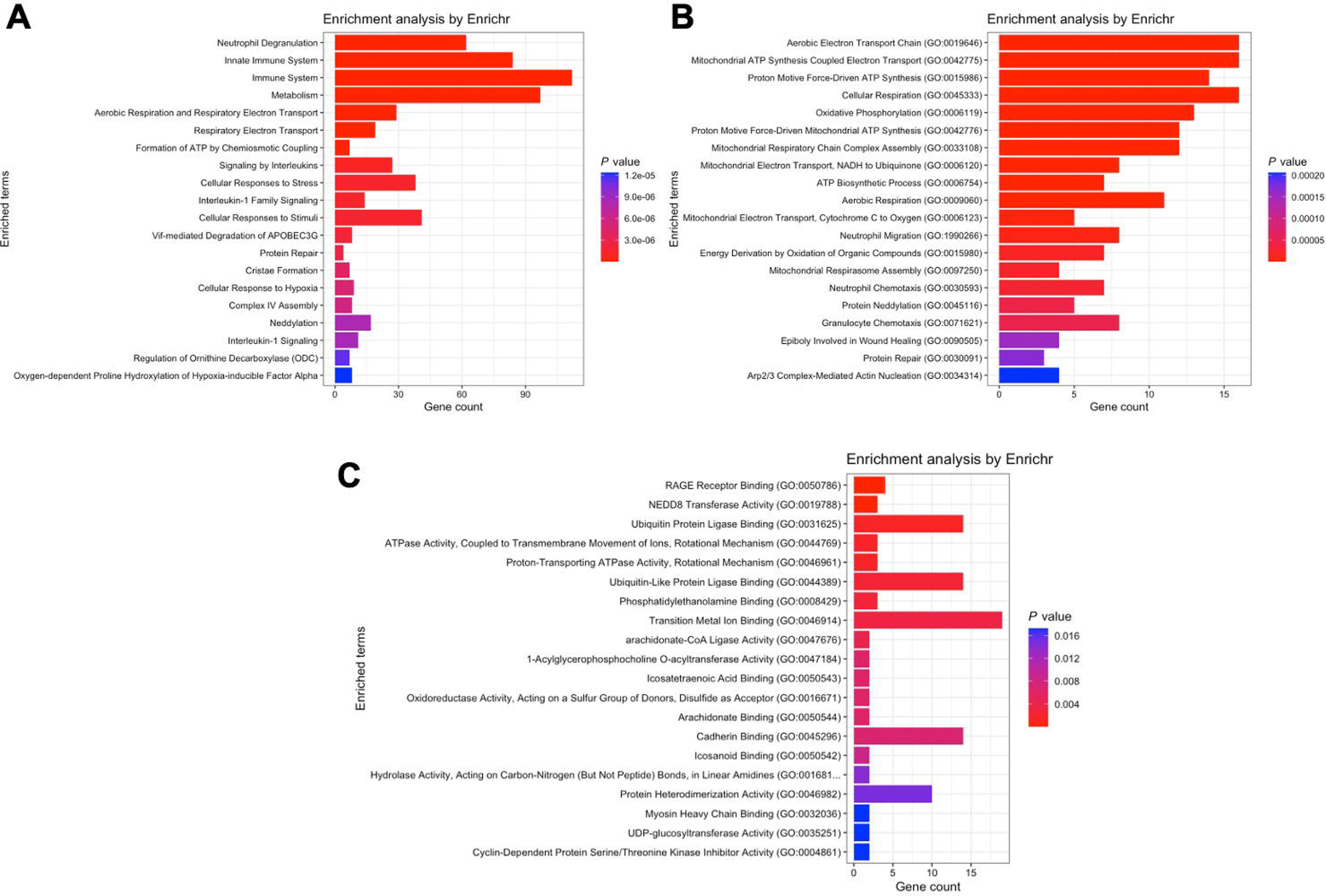
Pathway enrichment analysis for genes within module brown of the septic shock network. (A) Reactome pathway analysis indicating involvement in immune responses, metabolism, and respiratory electron transport. (B) GO biological processes associated with module brown, such as aerobic respiration and neutrophil chemotaxis. (C) GO molecular functions enriched in this module, including ubiquitin ligase binding and metal ion binding.

### COVID-19 gene network analysis parallels the sepsis network, confirming that S100A8 and S100A9 play a major role in ubiquitination and cellular respiration

**Figure 4A** exhibits the module-trait correlations of the COVID-19 network. Modules greenyellow and black are positively correlated with the COVID-19 state (Pearson coefficient equals 0.49 and 0.59, respectively). The expression profiles for modules greenyellow and black are shown in **Figures 4B and 4C**, respectively.

**Figure 4.**
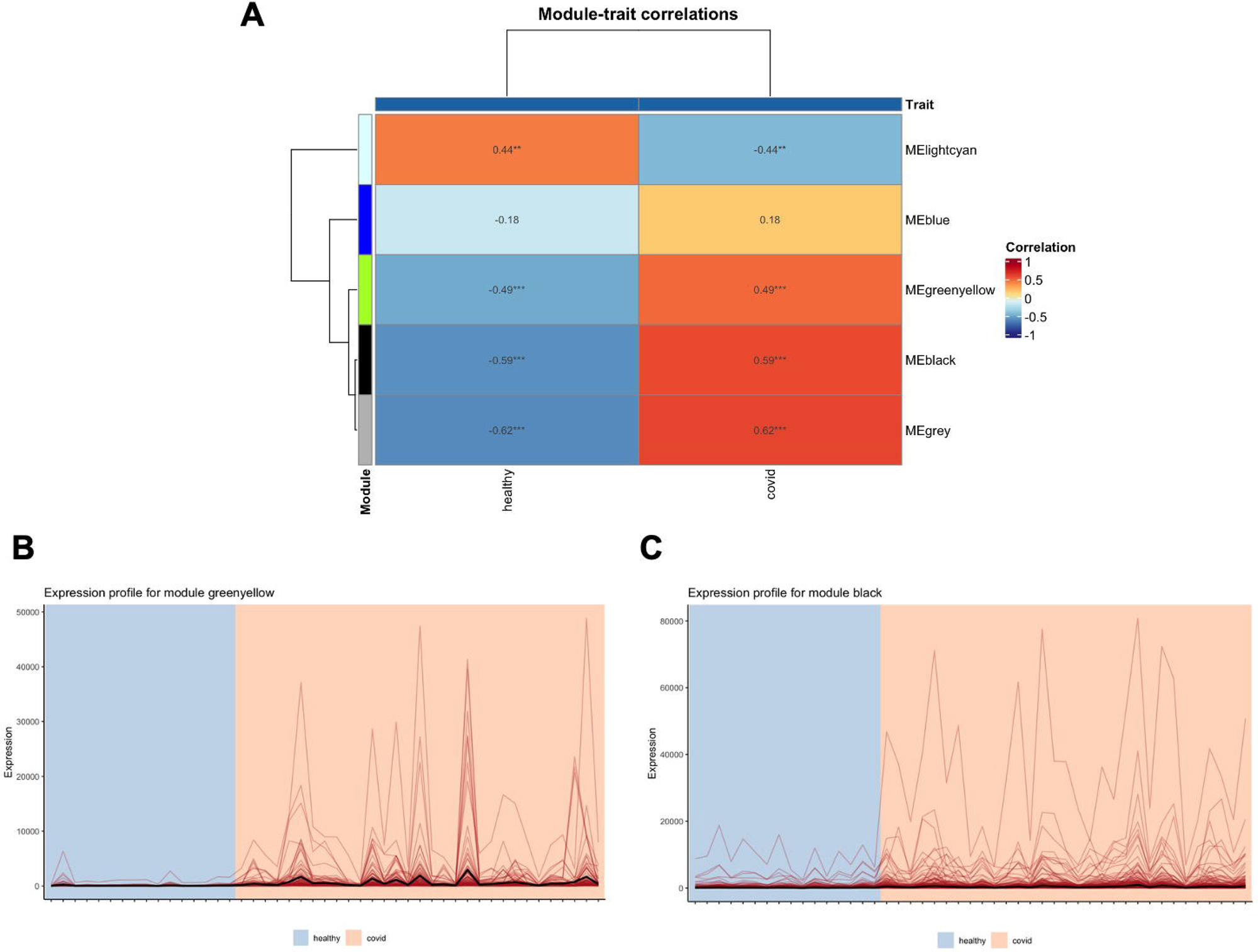
Module-trait relationship analysis of the COVID-19 network. (A) Correlation heatmap showing associations between module eigengenes (MEs) and COVID-19 status; positive correlations are depicted with warm colors. Expression profiles of (B) module greenyellow and (C) module black are shown as line plots across samples, with ME indicating module eigengenes.

Regarding module greenyellow, where *CAMP* and *DEFA1* to *DEFA4* are located, the Reactome database detected pathways such as binding and uptake by scavenger receptors, scavenging of heme, complement activation and FCGR activation (**Figure 5A**). The Gene ontology analyses, in turn, revealed pathways such as antimicrobial humoral response and defense to bacteria (biological processes, **Figure 5B**), as well as immunoglobulin receptor and iron binding (molecular functions, **Figure 5C**).

**Figure 5.**
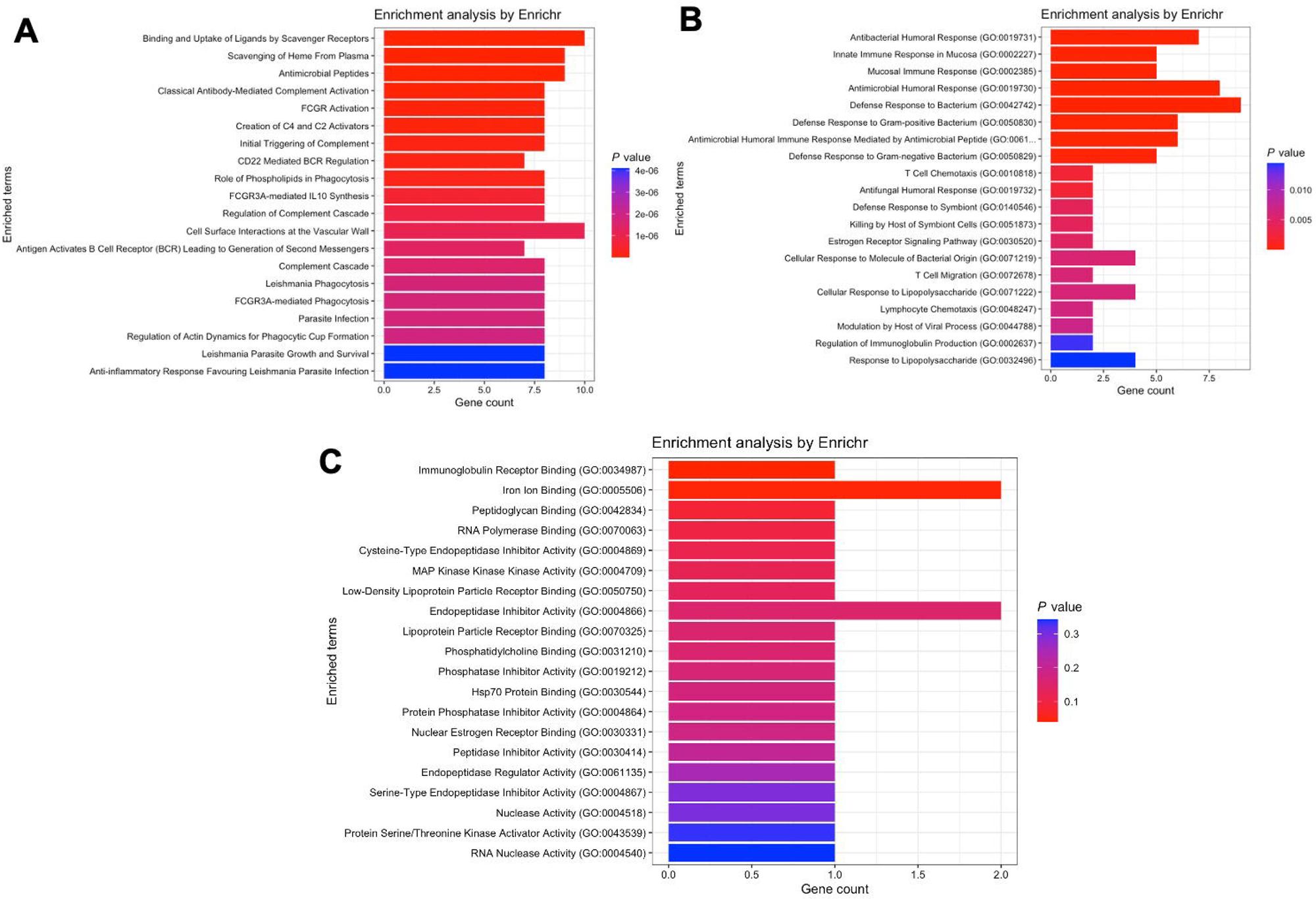
Pathway enrichment analysis for genes within module greenyellow of the COVID-19 network. (A) Reactome pathway analysis highlighting pathways such as scavenger receptor binding and uptake, heme scavenging, complement activation, and Fc gamma receptor activation. (B) GO biological processes related to antimicrobial humoral response and bacterial defense. (C) GO molecular functions including immunoglobulin receptor activity and iron binding.

The black module, however, which contains *S100A8* and *S100A9*, revealed results such as innate immunity activation (Reactome database, **Figure 6A**), mitochondrial electron transport and cellular respiration (Gene Ontology – biological processes, **Figure 6B**), ubiquitin ligase binding, cadherin binding and GTPase activity (Gene Ontology – molecular functions, **Figure 6C**).

**Figure 6.**
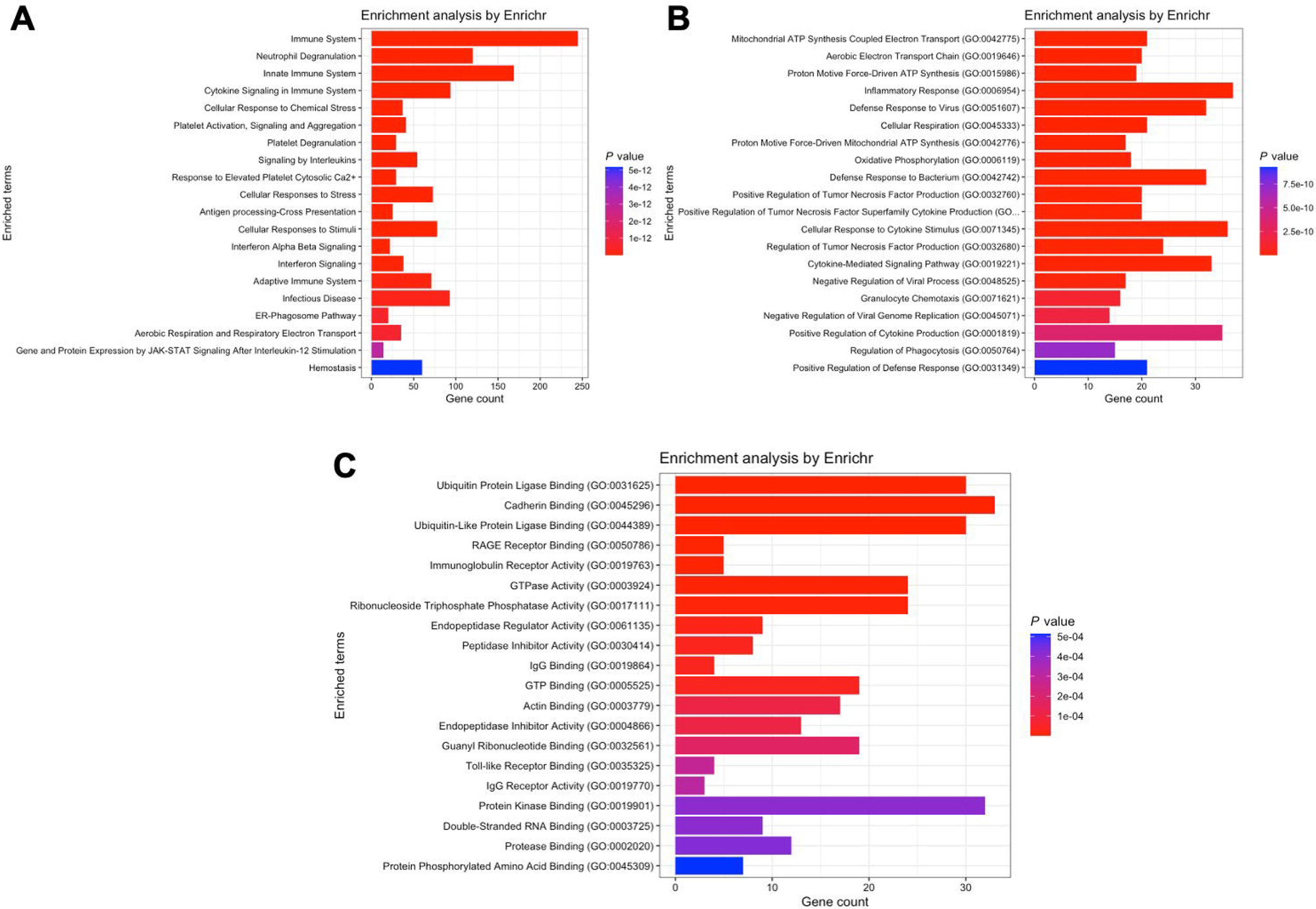
Pathway enrichment analysis for genes within module black of the COVID-19 network. (A) Reactome analysis indicating activation of innate immunity pathways. (B) GO biological processes emphasizing mitochondrial electron transport and cellular respiration. (C) GO molecular functions enriched in this module, such as ubiquitin ligase binding, cadherin binding, and GTPase activity.

### In the pancreatitis network, CAMP is in the same module as the S100A genes and alpha defensins are downregulated

**Figure 7A** shows the module-trait correlations of the acute pancreatitis network. Modules brown is positively correlated with the acute pancreatitis state (Pearson coefficient equals 0.77), but module blue is negatively correlated (Pearson coefficient equals -0.29). The expression profiles for modules blue and brown are shown in **Figures 7B and 7C**, respectively.

**Figure 7.**
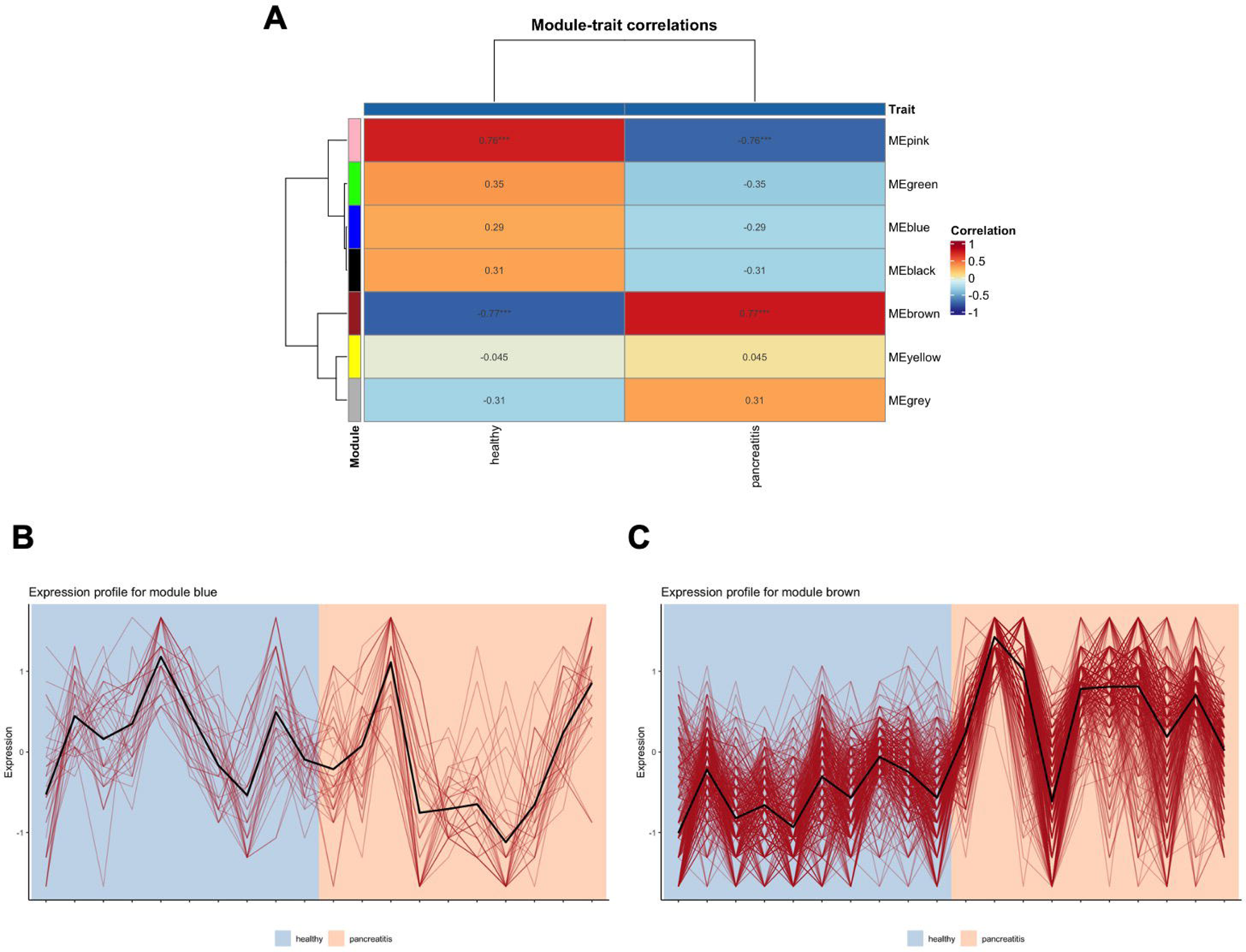
Module-trait relationship analysis of the acute pancreatitis network. (A) Correlation heatmap illustrating the association between module eigengenes (MEs) and pancreatitis status; positive correlation for module brown and negative correlation for module blue are shown. Expression profiles of (B) module blue and (C) module brown are displayed as line plots, with ME representing module eigengenes.

Module blue contains *DEFA1* to *DEFA3*. *DEFA4* was not detected in any module. Pathways analysis highlights results such as antimicrobial peptides (Reactome database, **Figure 8A**), mucosal immune response and steroid hormone signaling (Gene Ontology – biological processes, **Figure 8B**), as well as lncRNA and single-stranded DNA binding (Gene Ontology – molecular functions, **Figure 8C**).

**Figure 8.**
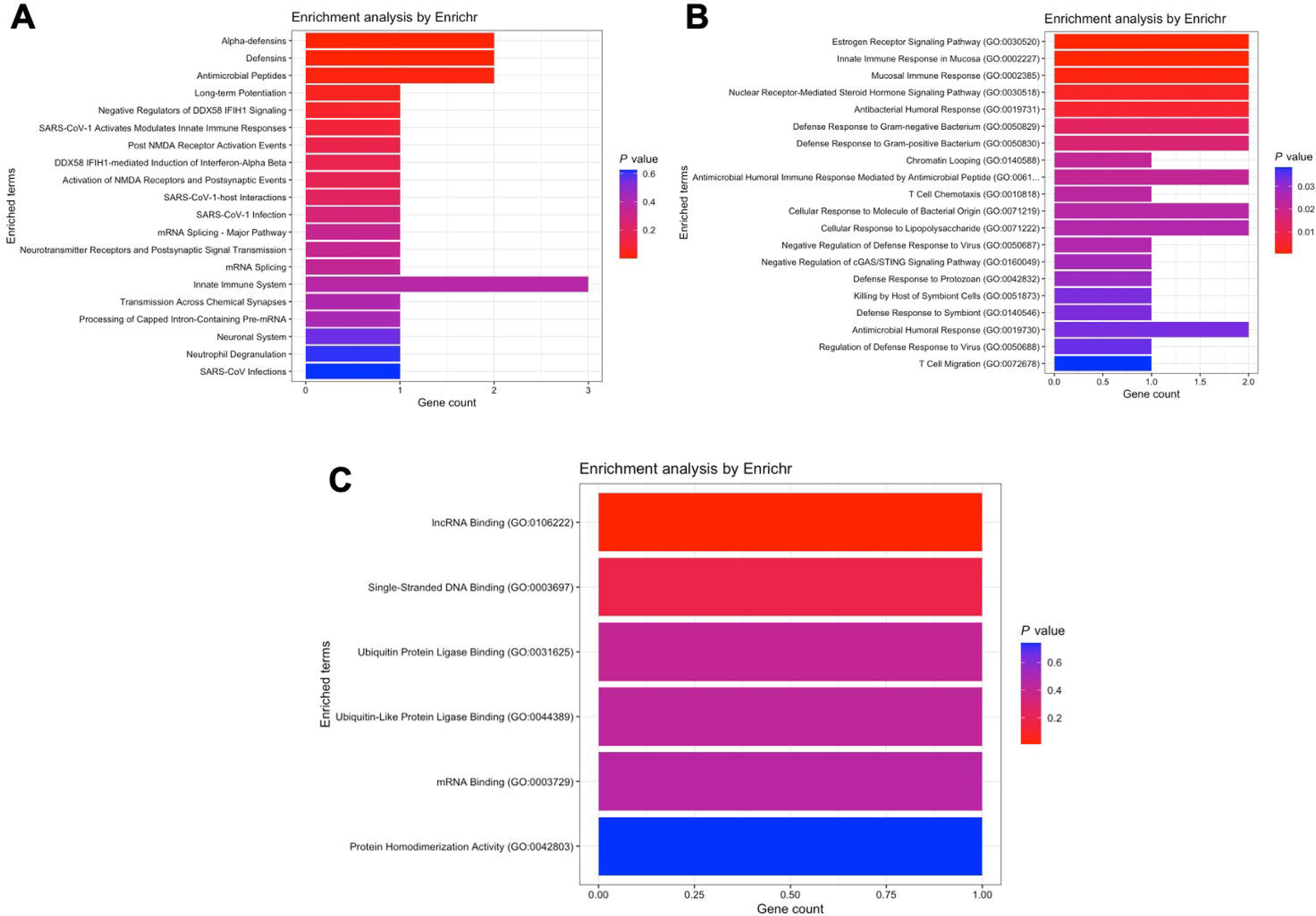
Pathway enrichment analysis for genes within module blue of the acute pancreatitis network. (A) Reactome pathway analysis highlighting antimicrobial peptides, mucosal immune responses, and steroid hormone signaling. (B) GO biological processes associated with this module include immune response and hormone signaling pathways. (C) GO molecular functions enriched, such as lncRNA and single-stranded DNA binding.

In module brown, surprisingly, *CAMP* appears together with *S100A8* and *S100A9*. The Reactome database analysis displays pathways such as innate and adaptive immune activation (**Figure 9A**), while the Gene Ontology databases reveal cytokines production and signaling (biological processes, **Figure 9B**), cadherin, ubiquitin ligase and protein kinase binding (molecular functions, **Figure 9C**).

**Figure 9.**
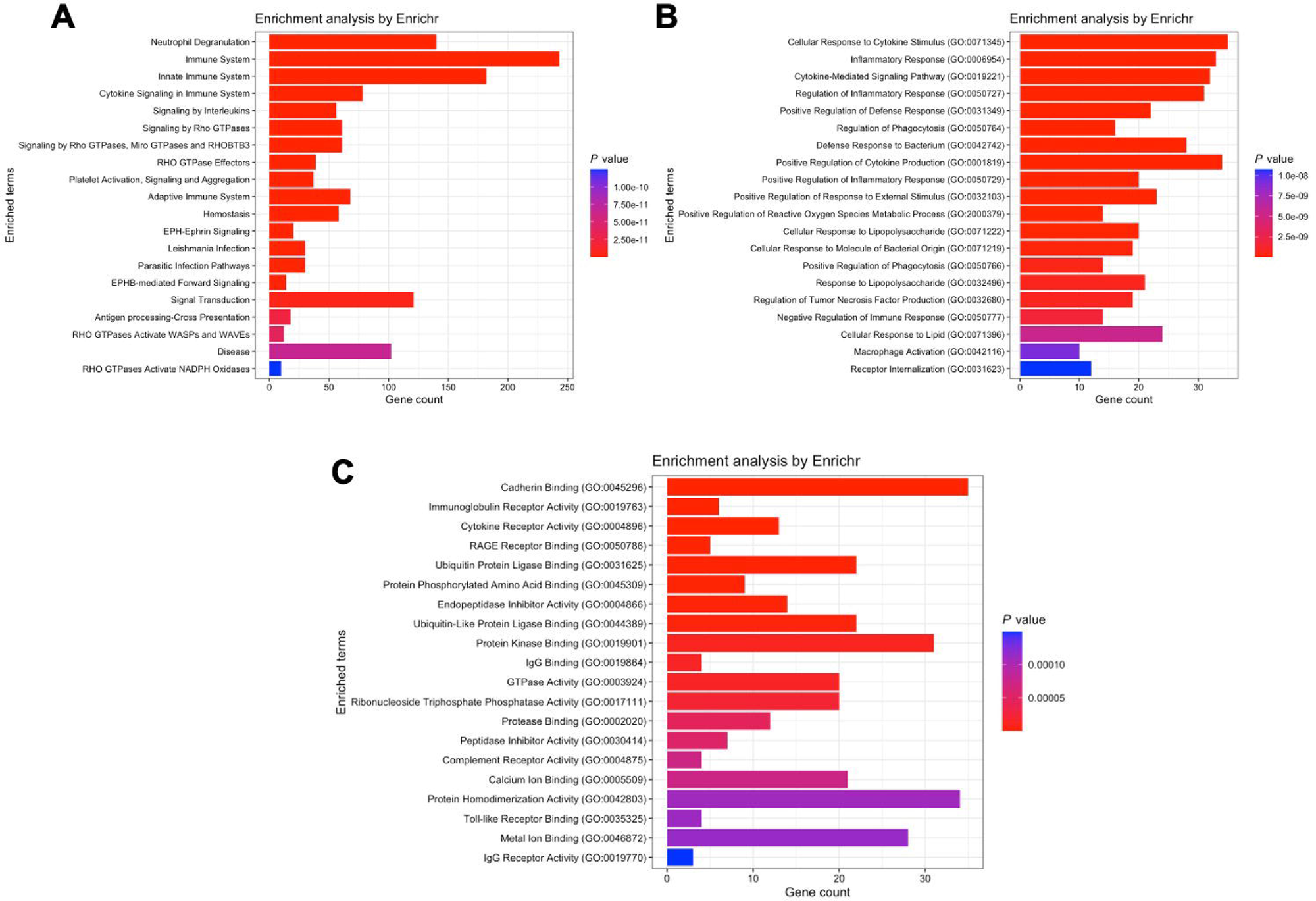
Pathway enrichment analysis for genes within module brown of the acute pancreatitis network. (A) Reactome pathways indicating innate and adaptive immune activation. (B) GO biological processes related to cytokine production and signaling. (C) GO molecular functions including cadherin binding, ubiquitin ligase binding, and protein kinase binding.

## 4. Discussion

Recent research investigating the role of antimicrobial peptides in *Drosophila* reveals that while defense against some pathogens relies on the activity of multiple AMPs, some AMPs exhibit a high degree of specificity, combating specific pathogens but not others [11, 12].

At the same time, several publications are putting in evidence that cathelicidins and defensins display a dual function, since depending on the underlying disease and the cellular microenvironment, they can induce both inflammatory or anti-inflammatory responses [3, 13, 14].

Here, we investigated the role of cathelicidin, alpha-defensins, S100A8, and S100A9 in systemic inflammation triggered by different insults, as we believe systemic inflammation is not a general process and antimicrobial peptides may act differently in each situation.

We found that during bacterial septic shock and severe COVID-19 infections, cathelicidin and alpha-defensins are activated together and act synergistically, participating in the innate host response to infection, even though the molecular functions detected in the analysis of the COVID-19 network (immunoglobulin receptor and iron binding) differed from those detected in the sepsis network (endopeptidase activity, calcium-dependent phospholipid binding, and protein kinase activity).

Regarding S100A8 and S100A9, their genes were also co-expressed in the presence of septic shock or severe COVID-19 infection, but in different modules than cathelicidin and alpha-defensins, unveiling distinct mechanisms of action. Indeed, analysis of the septic shock network suggests that calprotectin is part of the immune response but mainly participates in mechanisms related to mitochondria, chemotaxis, ubiquitin ligase, and metal ion binding pathways. Similarly, *S100A8* and *S100A9* were also associated with aerobic respiration and ubiquitin ligase binding in the context of COVID-19 infection.

S100A8 and S100A9 are important alarmins with a crucial role in the pathogenesis of septic shock [15] and COVID-19 infections [16]. They chelate metal nutrients (especially calcium and zinc), restricting access to essential metals necessary for pathogen growth and virulence factor activity. Calprotectin can also bind to several cell receptors, participating in various arms of host immunity [5]. Ubiquitin ligase binding, however, is an intriguing finding, since S100A8 and S100A9 have never been investigated in relation to the proteasome.

The role of calprotectin in the proteasome, however, has a rationale and deserves further investigation. Computational analysis of the human proteome has identified thousands of peptides with antimicrobial properties embedded in proteins with functions unrelated to the immune system [17, 18]. Proteases and proteasomal degradation could eventually liberate these antimicrobial peptides and allow them to exert activity. Indeed, a recent study describes that some products of proteasomal degradation possess antimicrobial properties, uncovering a previously unrecognized role for proteasomes in generating these peptides [19]. Along the same lines, degradation of alpha-defensins by proteases generated several fragments with different antimicrobial properties [20, 21]. I believe that it is possible, therefore, that calprotectin proteasomal degradation also leads to the generation of additional antimicrobial peptides.

Finally, we discuss the results of the acute pancreatitis network. They diverge from the sepsis and COVID-19 networks. Surprisingly, the *CAMP* gene is co-expressed with *S100A8* and *S100A9*, while *DEFA1* to *DEFA3* are located in a different module. *DEFA4* was not detected in any module. *CAMP*, *S100A8*, and *S100A9* are activated, when compared to healthy controls, but *DEFA1* to *DEFA3* gene expression is inhibited.

Regarding alpha-defensins, pathway analysis reveals inhibition of mucosal immune response, lncRNA, and single-stranded DNA binding. Pathway analysis of the module containing *CAMP*, *S100A8*, and *S100A9*, however, shows pathway enrichment such as innate and adaptive immunity activation, cytokine signaling, ubiquitin ligase, and protein kinase binding. From our point of view, these results support our hypothesis that systemic inflammation is a finely tuned response to extensive insults and that antimicrobial peptides operate in conjunction in this scenario, but not always synergistically or through the same mode of action.

This study has a limitation, it was done using whole blood. Future studies should focus on single cells RNA-sequencing and investigate not only the blood, but also organs such as the brain, lungs, kidneys and the gut. Further exploration into the cellular sources and context-dependent functions of these antimicrobial peptides may elucidate their precise roles in modulating inflammation. I believe that heterogeneous expression patterns of AMPs can be found within different cell populations and tissues, suggesting that their activity could be highly localized and influenced by cell type and microenvironment. Understanding the cellular and molecular determinants that dictate whether these peptides promote or suppress inflammation will be crucial for developing targeted therapeutic strategies, especially in complex syndromes like sepsis and COVID-19 where immune dysregulation is prevalent.

Moreover, the potential therapeutic implications of modulating AMP activity warrant further investigation. Given their dual roles in inflammation and antimicrobial defense, strategies aimed at enhancing beneficial functions or inhibiting detrimental effects could improve patient outcomes. A deeper understanding of the regulation and activity of these peptides, particularly in specific tissues or disease stages, could facilitate the development of novel interventions capable of fine-tuning immune responses in systemic inflammation.

## 5. Conclusion and Hypothesis

Our study demonstrates that antimicrobial peptides - specifically cathelicidin, alpha-defensins, and the S100 proteins S100A8 and S100A9 - play distinct and context-dependent roles in systemic inflammation triggered by different pathological insults. In bacterial septic shock and severe COVID-19, cathelicidin and alpha-defensins are co-activated and synergistically contribute to the innate immune response, while *S100A8* and *S100A9* engage in separate pathways related to mitochondrial function and ubiquitin ligase activity. Conversely, in acute pancreatitis, these peptides exhibit a divergent expression pattern, with *CAMP* co-expressed alongside S100 genes and alpha-defensins downregulated, suggesting differing immune modulation mechanisms. These findings emphasize that systemic inflammation is a finely tuned, multifaceted response where antimicrobial peptides act in coordination but through varied modes of action depending on the disease context. Further investigation into their mechanistic roles, including potential involvement in proteasomal processing, may reveal novel therapeutic targets for managing critical illnesses characterized by systemic inflammation.

## Data Availability

All data produced in the present study are available upon reasonable request to the authors

## Author contributions

FPS conceived the study, performed the computational analyses and wrote the article.

## Acknowledgements

English proofreading was performed by chatGPT.

